# GWASHub: An Automated Cloud-Based Platform for Genome-Wide Association Study Meta-Analysis

**DOI:** 10.1101/2025.10.21.25338463

**Authors:** Nicholas Sunderland, Drew Hite, Patrick Smadbeck, Quy Hoang, Dong-Keun Jang, Vinicius Tragante, Jiayue-Clara Jiang, Sonia Shah, Lavinia Paternoster, Noel P. Burtt, Jason Flannick, R Thomas Lumbers, the HERMES Consortium & Cardiovascular Knowledge Portal

## Abstract

Genome-wide association studies (GWAS) often aggregate data from millions of participants across multiple cohorts using meta-analysis to maximise power for genetic discovery. The increase in availability of genomic biobanks, together with a growing focus on phenotypic subgroups, genetic diversity, and sex-stratified analyses, has led GWAS meta-analyses to routinely produce hundreds of summary statistic files accompanied by detailed meta-data. Scalable infrastructures for data handling, quality control (QC), and meta-analysis workflows are essential to prevent errors, ensure reproducibility, and reduce the burden on researchers, allowing them to focus on downstream research and clinical translation. To address this need, we developed GWASHub, a secure cloud-based platform designed for the curation, processing and meta-analysis of GWAS summary statistics.

GWASHub features i) private and secure project spaces, ii) automated file harmonisation and data validation, iii) GWAS meta-data capture, iv) customisable variant QC, v) GWAS meta-analysis, vi) analysis reporting and visualisation, and vii) results download. Users interact with the portal via an intuitive web interface built on Nuxt.js, a high-performance JavaScript framework. Data is securely managed through an Amazon Web Services (AWS) MySQL database and S3 block storage. Analysis jobs are distributed to AWS compute resources in a scalable fashion. The QC dashboard presents tabular and graphical QC outputs allowing manual review of individual datasets. Those passing QC are made available to the meta-analysis module. Individual datasets and meta-analysis results are available for download by project users with appropriate access permissions. In GWASHub, a “project” serves as a virtual workspace spanning an entire consortium, allowing individuals with different roles, such as data contributors (users) and project coordinators (main analysts), to collaborate securely under a unified framework. GWASHub has a flexible architecture to allow for ongoing development and incorporation of alternative quality control or meta-analysis procedures, to meet the specific needs of researchers. GWASHub was developed as a joint initiative by the HERMES Consortium and the Cardiovascular Knowledge Portal, and access to the platform is free and available upon request.

GWASHub addresses a critical need in the genetics research community by providing a scalable, secure, and user-friendly platform for managing the complexity of large-scale GWAS meta-analyses. As the volume and diversity of GWAS data continue to grow, platforms like GWASHub may help to accelerate insights into the genetic architecture of complex traits.

## 1. Introduction

Genome-wide association studies (GWAS) have drastically increased our understanding of the genetic basis for complex diseases. Detecting the small additive effects from millions of variants across the genome requires large sample sizes, which are typically achieved through meta-analysis.^1–3^ In large-scale studies, this routinely involves managing hundreds of data files, a number that grows significantly when stratifying by sex or genetic ancestry. Effective data handling strategies and quality control procedures are essential to prevent inadvertent errors, ensure reproducibility, and reduce the time burden on researchers.

GWAS consortia analysts typically develop their own quality control (QC) procedures, many of which have been published as software packages, for example EasyQC^5^ and BIGwas^6^. While these resources help streamline aspects of GWAS analysis, critical friction points remain throughout the research workflow. Aggregating GWAS data files is still largely manual, and issues such as inconsistent file naming, variable data format specifications, and incomplete or mismatched meta-data are common. QC procedures rely on access to large, standardised reference samples. Although many of these resources are publicly available, their integration introduces additional data formats and complexity into analysis pipelines. Additionally, large meta-analysis projects typically require access to high-performance computing environments, with adequate memory and support for parallel processing. Multiple different GWAS meta-analysis software options are freely available, each with their own strengths and weaknesses, for example fixed and random effects models, application of genomic control, and handling multi-ancestry analyses.^7,8^ Input formats and runtime parameters differ between software adding further complexity if multiple meta-analyses are to be run. Cloud-based data infrastructures offer an alternative, scalable solution for coordination of data contributions and analyses, increasing transparency and accessibility of data within the consortium.

Here we present the online platform GWASHub to address these issues, with private data repository and project spaces, customisable for individual consortia requirements. We demonstrate the utility of this resource by replicating a recent GWAS meta-analysis of dilated cardiomyopathy (DCM).

## 2. Design and Implementation

### Consortium context and workflow

In a typical GWAS consortium such as HERMES, multiple cohorts contribute genetic and phenotypic data to a collaborative research effort. Each consortium includes data contributors, who generate GWAS summary statistics and phenotypic meta-data for their cohort, and main analysts, who perform additional QC procedures to ensure consistency across studies. Coordinating analysts then carry out meta-analyses and downstream post-GWAS analyses. GWASHub mirrors these real-world roles and workflows through project spaces, user accounts with defined roles, and versioned datasets and meta-analyses, providing a digital framework that reduces the organisational burden on the consortium.

### Portal oOverview

The GWAS portal is a secure cloud-based platform designed for the curation, processing and analysis of GWAS summary statistics within private project spaces. The portal implements automated file validation and standardisation procedures through an intuitive web interface, followed by a customisable QC module. The QC dashboard presents tabular and graphical quality control outputs allowing manual review of individual datasets. Datasets passing QC are made available to the meta-analysis module. Individual datasets and meta-analysis results are available for download by project users with appropriate access permissions.

### Frontend user interface

The web interface is built on Nuxt.js, a high performance Javascript framework enabling fast page loading and seamless user interactions. The frontend communicates with the backend via a RESTful API exposed by a FastAPI Python server.

### Backend API and computations workflow

The backend FastAPI Python server runs on an Amazon Web Services (AWS) EC2 instance. The API ensures secure access through token-based authentication. Dataset and meta-data management is handled using a MySQL database running in the AWS Relational Database Service (RDS). Job scheduling of QC and meta-analyses is facilitated by batch processing in both AWS Batch and AWS Elastic MapReduce (EMR) environments, respectively, enabling execution on scalable cloud resources. Jobs read their input datasets from AWS S3 block storage and write out result files, processing summaries and figures back to the block storage. Versioned workflows ensure reproducibility of results.

### Workflows

At project conception a private portal space is created and user accounts created with either ‘uploader’ (study analyst) and/or ‘reviewer’ (main analyst) permissions. Users interact with several dashboards providing upload, QC, and meta-analysis features. An overview of the portal architecture and analysis modules is presented in **Figure 1**.

**Figure 1.**
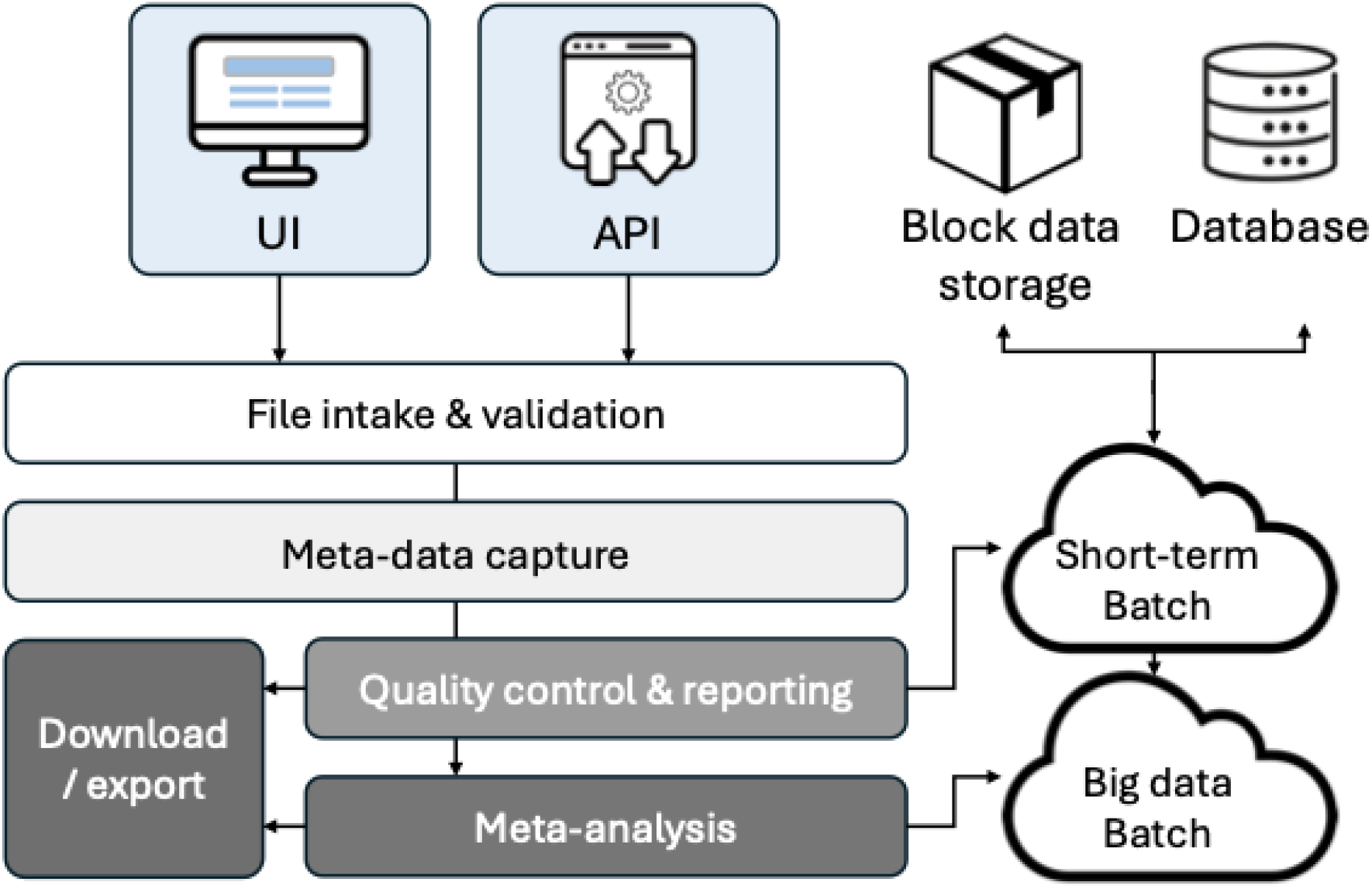
Portal GWAS Meta-analysis Pipeline.

#### File upload

On GWAS file upload, the portal implements basic file validation checks and provides informative error messages to the user if these fail (**Supplementary Table 1 & Supplementary Figure 1A**). Expected data types and value ranges are enforced - this validation helps catch high-level common errors such as missing data, incorrectly formatted columns, or inappropriate values. Early file validation prevents irrecoverable errors propagating, whilst the study analyst is engaged with the project and able to rectify the issues. In addition, a minimal set of mandatory meta-data fields is collected at file upload, ensuring the population under study and the methods used to generate the dataset are adequately described (**Supplementary Table 2 & Supplementary Figures 1B-F**).

#### Quality control

After successful file upload, a QC job is automatically scheduled to be run on the cloud platform. The QC steps are presented in **Supplementary Table 3**. Briefly, variants are harmonised to a reference genome assembly (Homo_sapiens.GRCh37.dna.primary_assembly.fa.gz), strand flipping is attempted for non-matching variants and those that remain unharmonised are flagged for removal. A summary table of harmonised variant counts and number of allele flips is presented along with examples of variants that have been removed from the analysis (**Supplementary Figure 2B**). Reported study effect allele frequencies are compared to ancestry-specific reference files from the 1000 Genomes Project for generation of a comparative allele frequency plot. Although no variant is excluded based on allele frequency deviation from the reference file by default, this can be implemented using the adjustable QC parameters. A PZ plot is generated based on the reported P-value and that determined from the Z-test derived from the reported beta and standard error. Finally, a Manhattan plot and QQ plot is presented for each dataset (**Supplementary Figure 2C**). After completion, the uploader, as well as the main project analyst, are able to review the QC results in the dataset explorer window. A dataset download link is provided to the lead analyst in case more refined exploration of the dataset is required. Once satisfactory QC has been confirmed, the lead analysis is required to flag the QC process as complete and satisfactory, using a button on the user interface, making it available for import into the meta-analysis module.

#### Meta-analysis

The meta-analysis dashboard displays all datasets having passed QC. Users with ‘reviewer’ permissions are able to submit meta-analysis jobs by selecting the desired input datasets and runtime parameters (**Supplementary Figure 3A**). The default meta-analysis pipeline runs an inverse-variance weighted meta-analysis with fixed-effects. However, this can be configured at project instantiation to use any standard meta-analysis software required by the project.

#### Visualisation

Once complete, the meta-analysis results file is made available for download alongside summary Manhattan and QQ plots (**Supplementary Figure 3B**).

#### GitHub repository

This repository provides a walk-through demonstration and detailed documentation on the GWASHub (https://github.com/broadinstitute/data-registry). We welcome user feedback and feature requests; please submit these through the GitHub repository.

## 3. Performance evaluation

We evaluated the performance of the modules’ execution time across realistic GWAS meta-analysis projects of increasing scale (**Table 1**). File upload and validation time per dataset ranged from 7 seconds for a dataset containing 2 million variants, to 32 seconds for 20 million variants. The QC module execution time ranged from 6 minutes for a dataset of 2 million variants, to 17 minutes for 20 million variants. The meta-analysis module execution time ranged from 1.6 hours for 5 datasets of 2 million variants each, to 9.1 hours for 20 datasets of 20 million variants each.

**Table 1.**
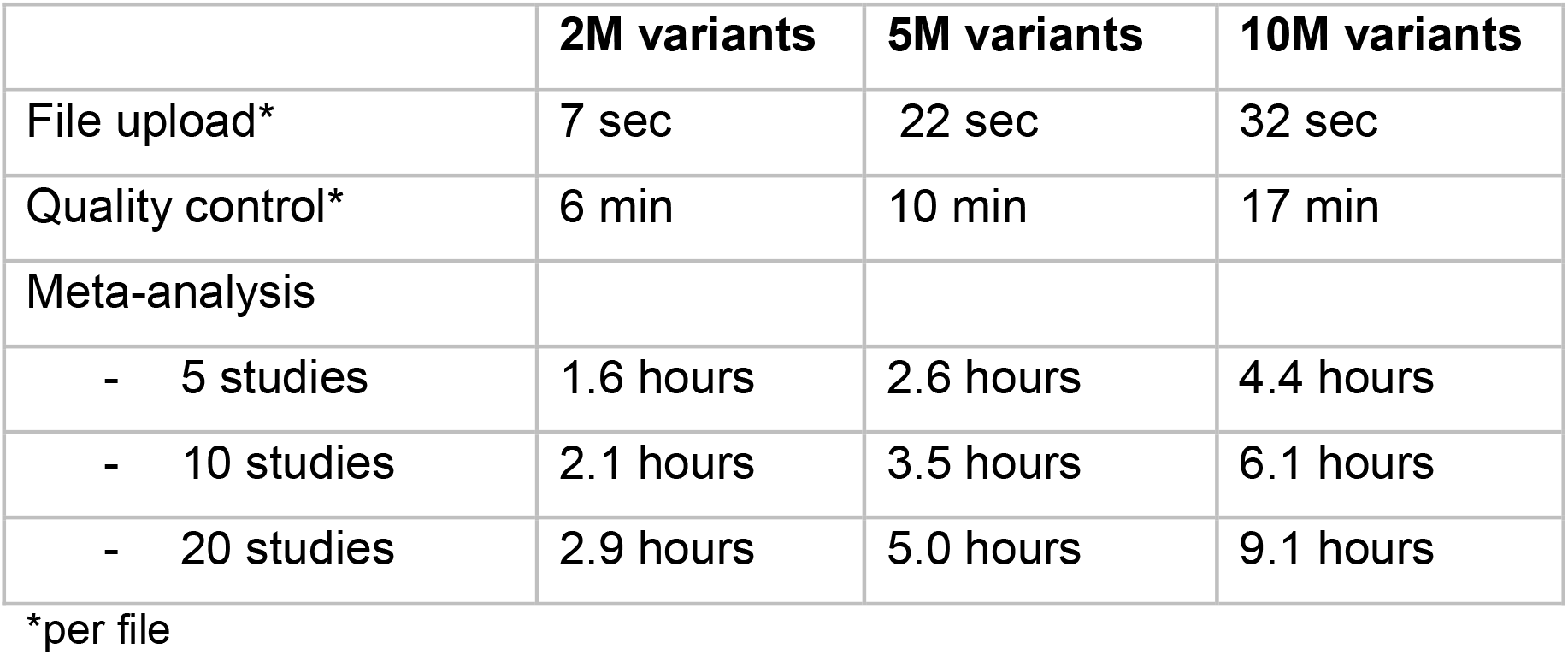
Execution time for GWAS projects by sample size.

Although the end-to-end workflow time will exceed the sum of the individual module runtimes, most of this additional time reflects analyst activities such as meta-data entry and QC output review. A smaller overhead arises from provisioning and initialising cloud resources for each analysis run, though this is typically minimal.

Finally, we replicated a meta-analysis of dilated cardiomyopathy case-control GWAS summary statistics using GWAMA, confirming the expected effect estimates of the meta-analysed variants through the GWASHub portal pipeline (**Supplementary Figure 4**).

## 4. Discussion

GWASHub provides an effective and scalable solution to running GWAS meta-analyses, overcoming many of the issues faced by analysts involved in these projects. Private project spaces offer secure and customisable environments in which to run analyses with rapid acquisition of results along with key quality control report elements. The pipeline substantially streamlines processing compared to manual file curation, data checking, and meta-analysis setup, improving efficiency and reducing the potential for human error.

The platform eliminates the need for time consuming file reformatting and the numerous email exchanges that are typically required to obtain meta-data from studies participating in GWAS meta-analyses consortia. In addition, the platform maintains an aggregate dataset to which new GWAS studies can be added as they become available with minimal overhead. It also supports flexible generation of leave-one-out meta-analyses to enable, for example, evaluation of polygenic scores, or the conduct of two sample Mendelian randomisation studies.

GWASHub is a platform that reduces the burden of routine data handling and analysis whilst removing the potential for human error. It is aimed at supporting large-scale studies and providing genetics researchers more time to focus on impactful downstream scientific questions.

For further information or to discuss potential use of GWASHub, interested researchers are encouraged to contact Tom Lumbers or Jason Flannick, or get in touch through the GWASHub GitHub repository (https://github.com/broadinstitute/data-registry).

We welcome user feedback and feature requests. Please submit these through the Portal GitHub repository issues board This repository also includes a walk-through demonstration and detailed documentation.

## Conflicts of interest

None

### Acknowledgements

NS is funded by the GW4-CAT Wellcome PhD programme. R.T.L. is supported by British Heart Foundation Special Project Grant SP/F/24/150066, National Institute for Health Research University College London Hospitals Biomedical Research Centre (NIHR203328), National Institutes of Health 5R01HL167509-02, and Medical Research Council Rare Disease Research UK Cardiovascular Initiative MR/Y008235/1.

## Data availability

The GWAS portal software is openly available on Github: https://github.com/nicksunderland/data-registry-api (backend API) and https://github.com/broadinstitute/data-registry (frontend)

