## Supplemental Figures for "GWASHub: An Automated Cloud-Based Platform for Genome-Wide Association Study Meta-Analysis"

**Supplementary figures**

**Figure 1A - GWAS dataset upload and column mapping**

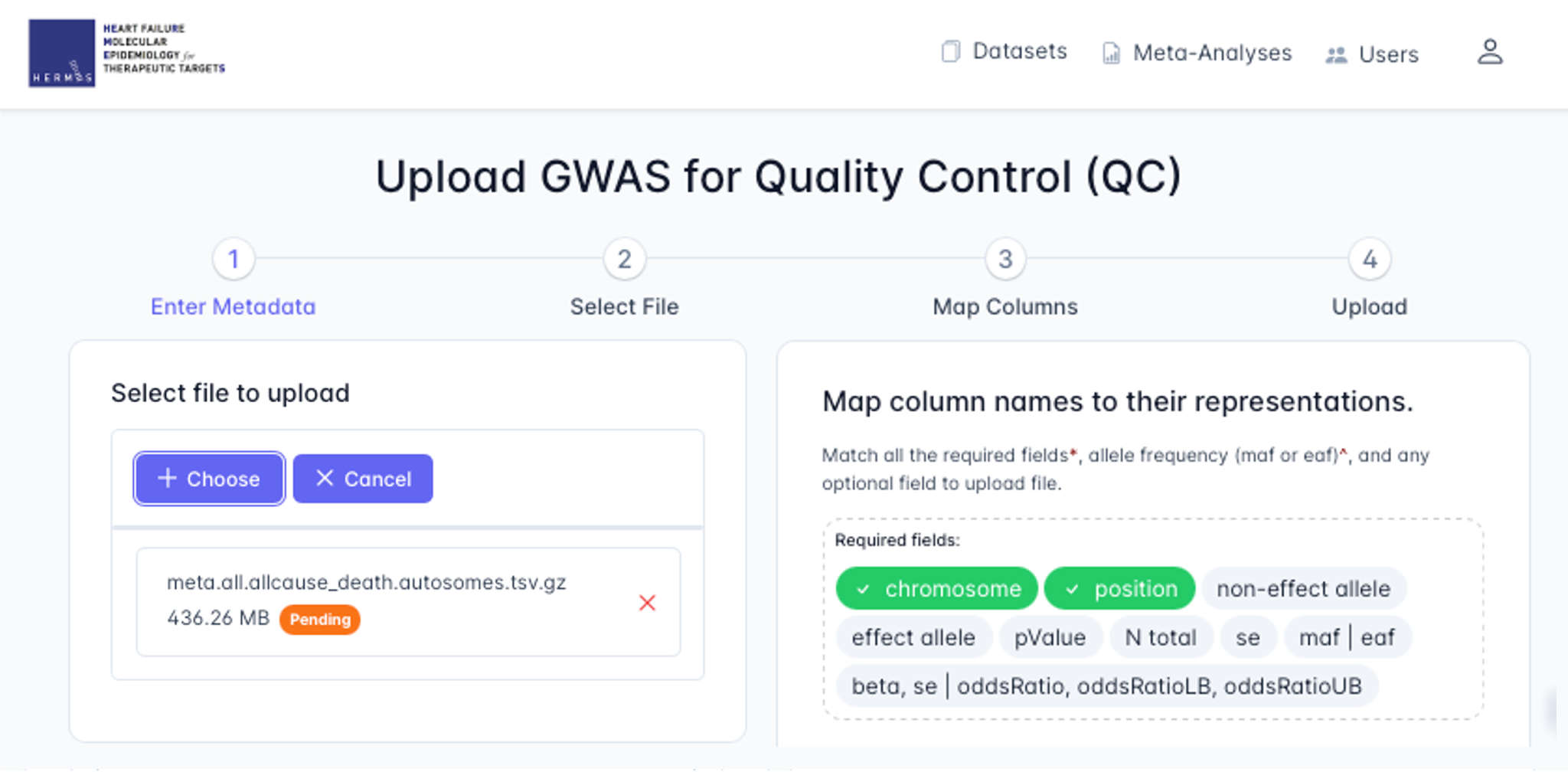

**Figure 1B - study and file metadata input**

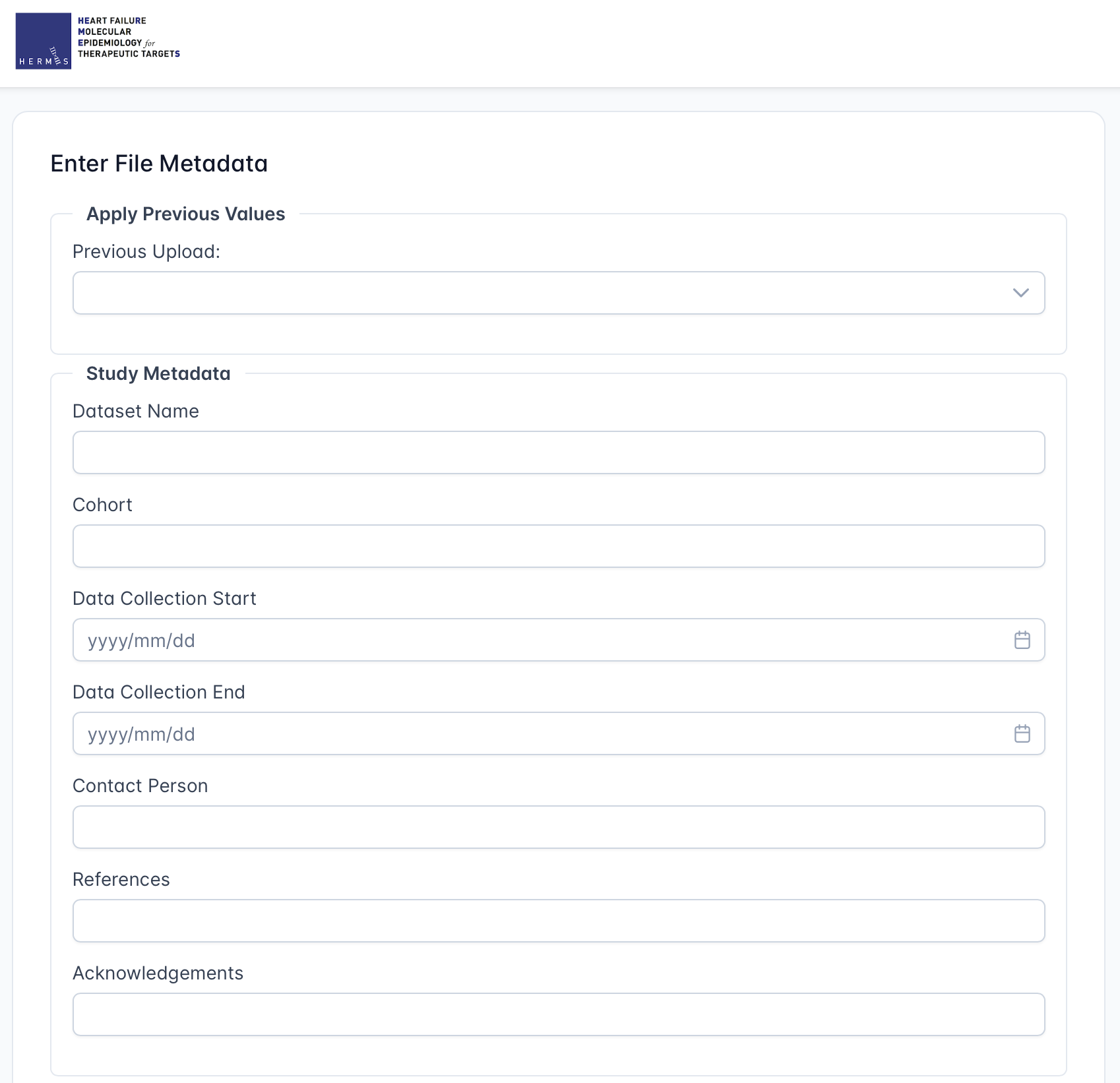

**Figure 1C - participants metadata input**

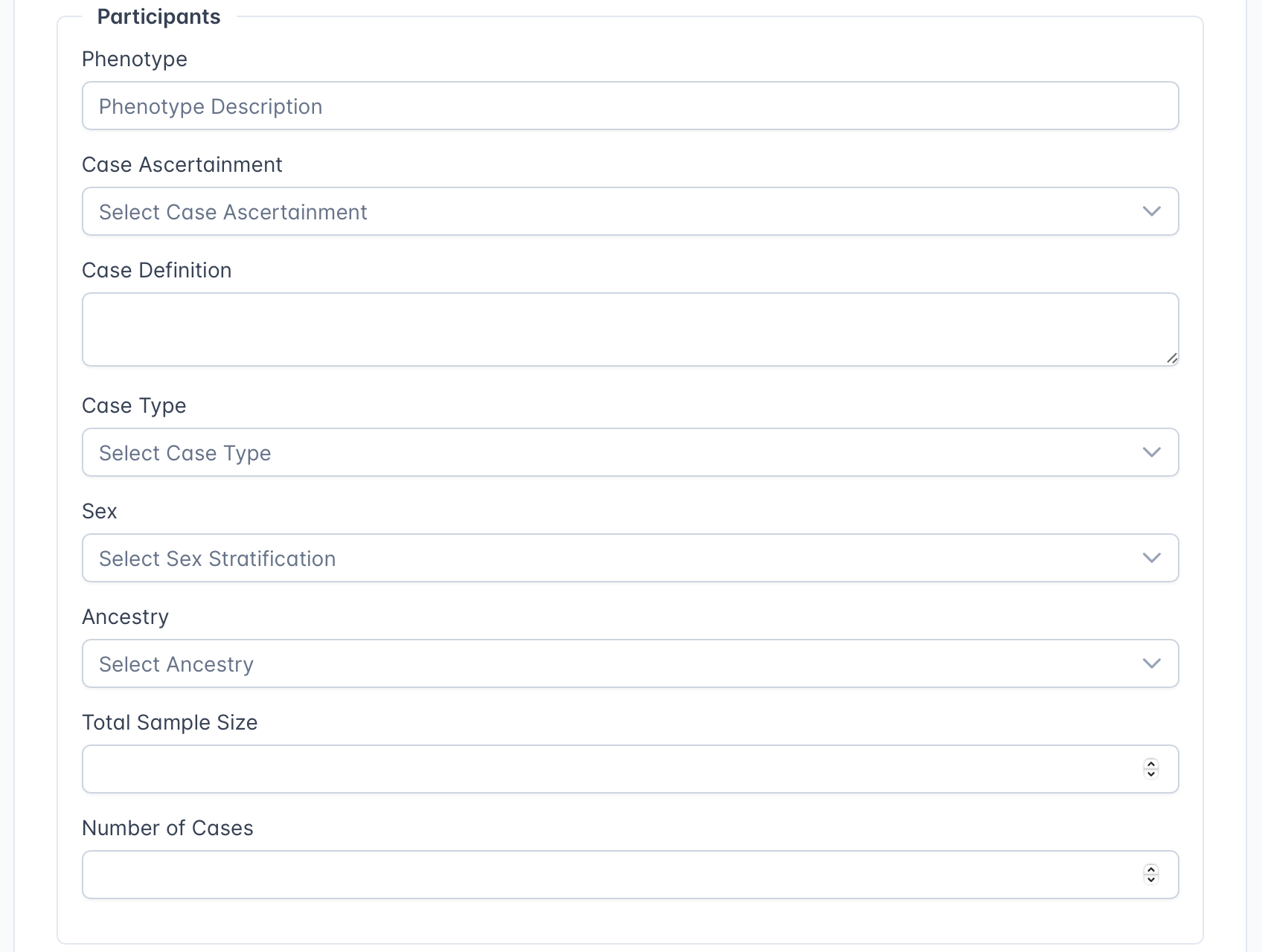

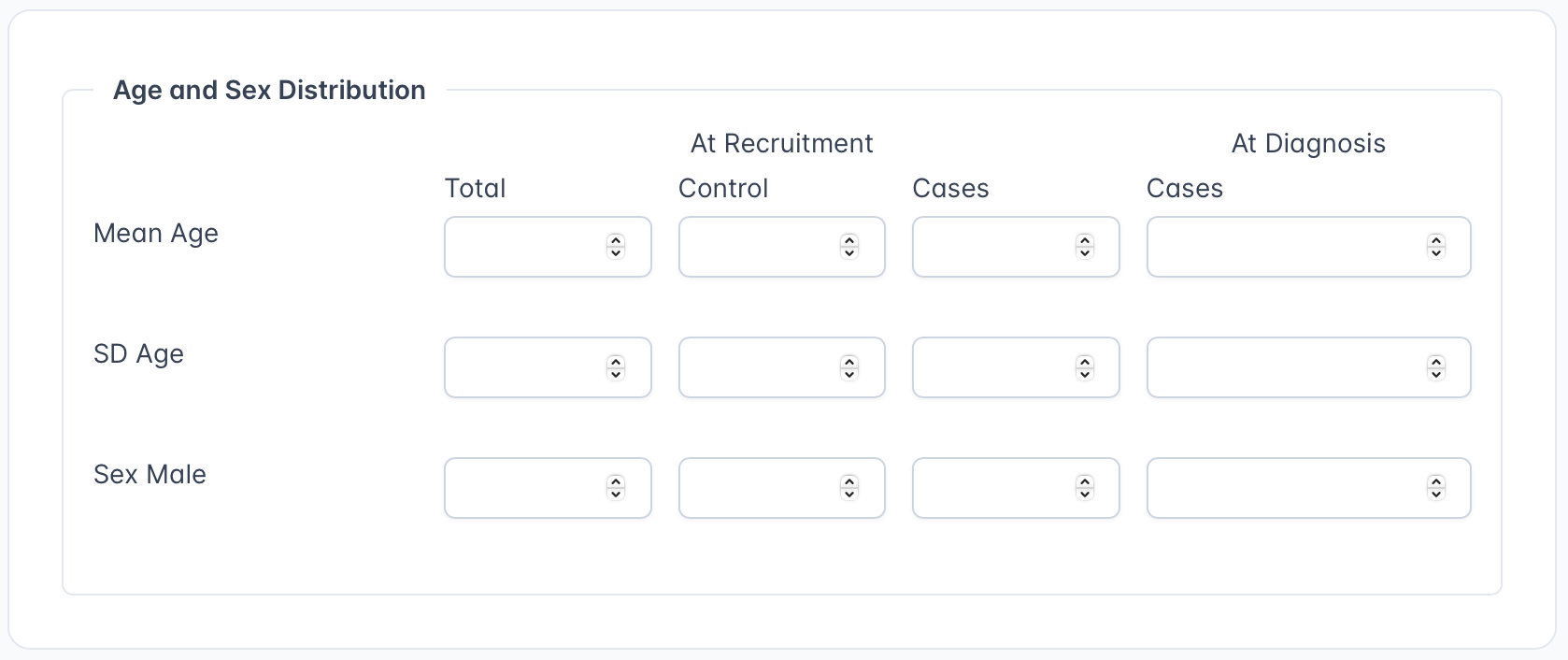

**Figure 1D - genotyping metadata input**

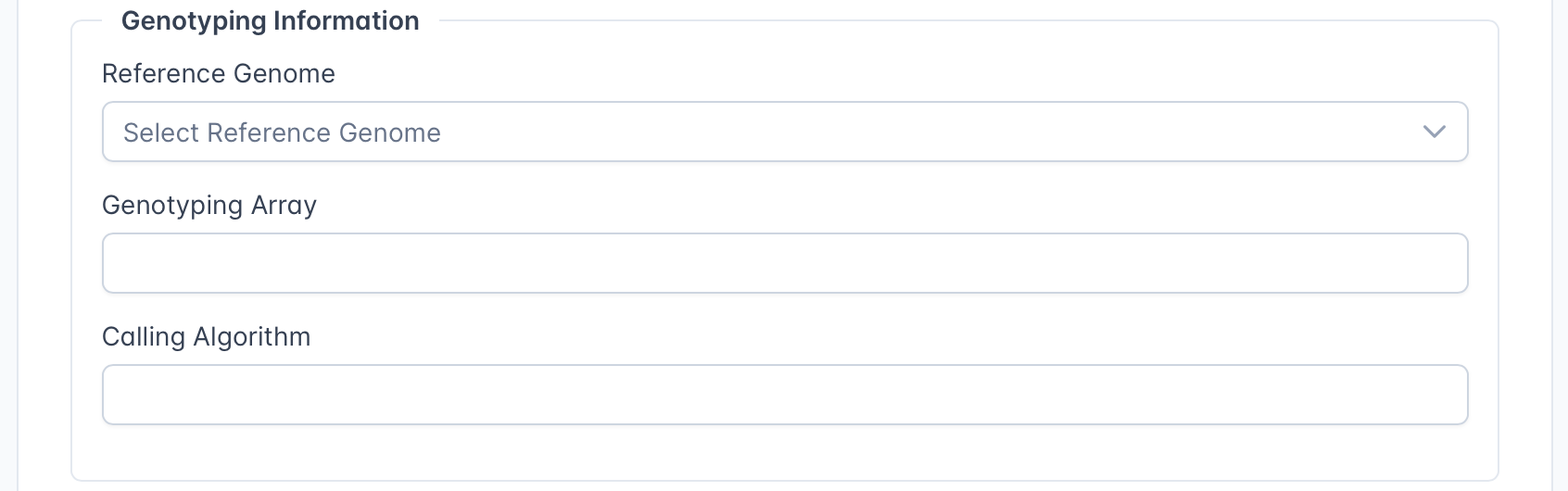

**Figure 1E - imputation metadata input**

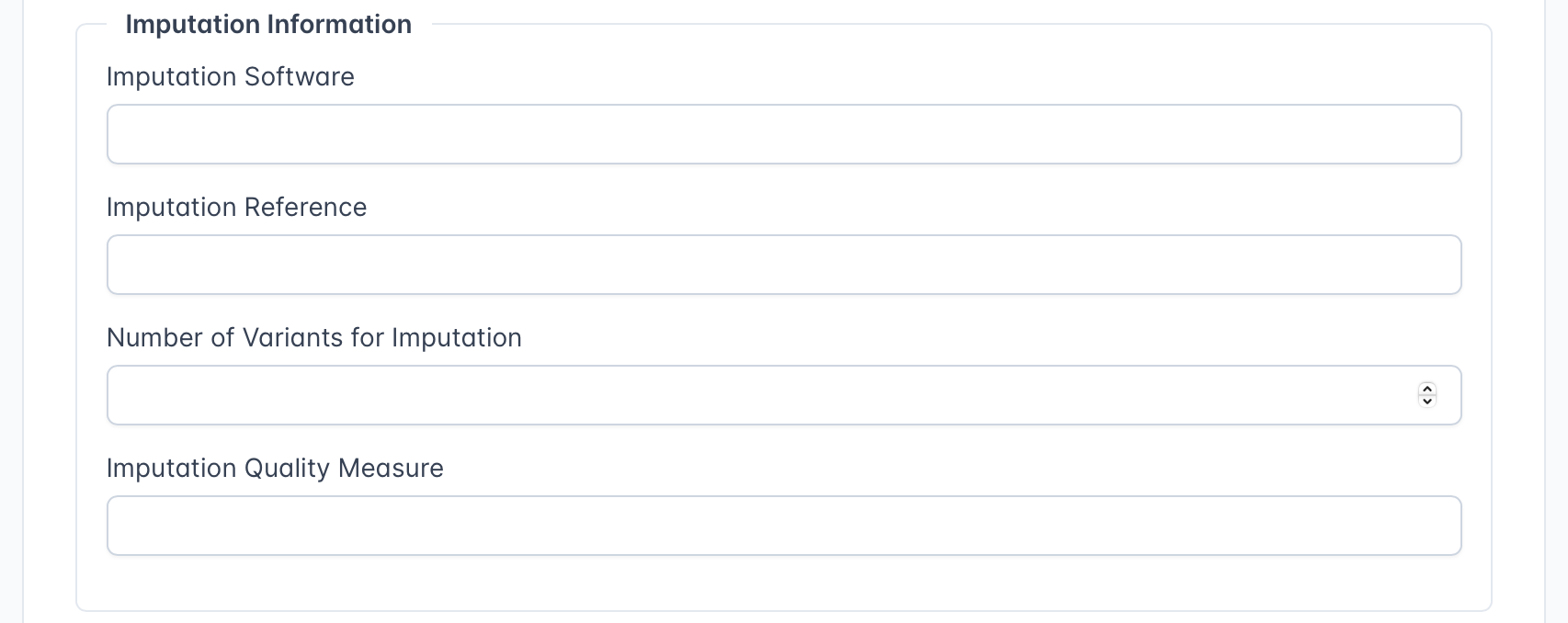

**Figure 1F - GWAS quality control metadata input**

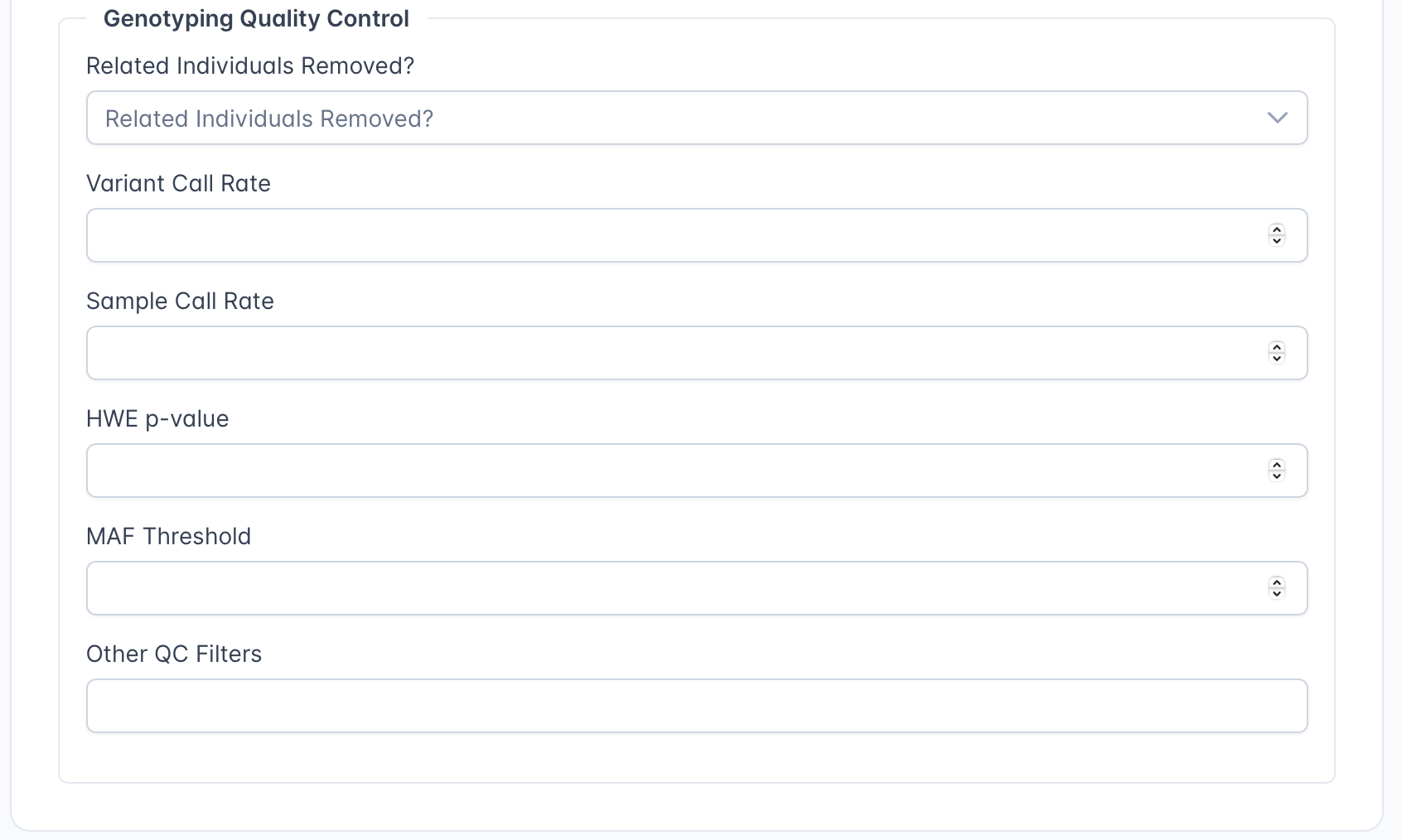

**Figure 2A - GWAS quality control dashboard**

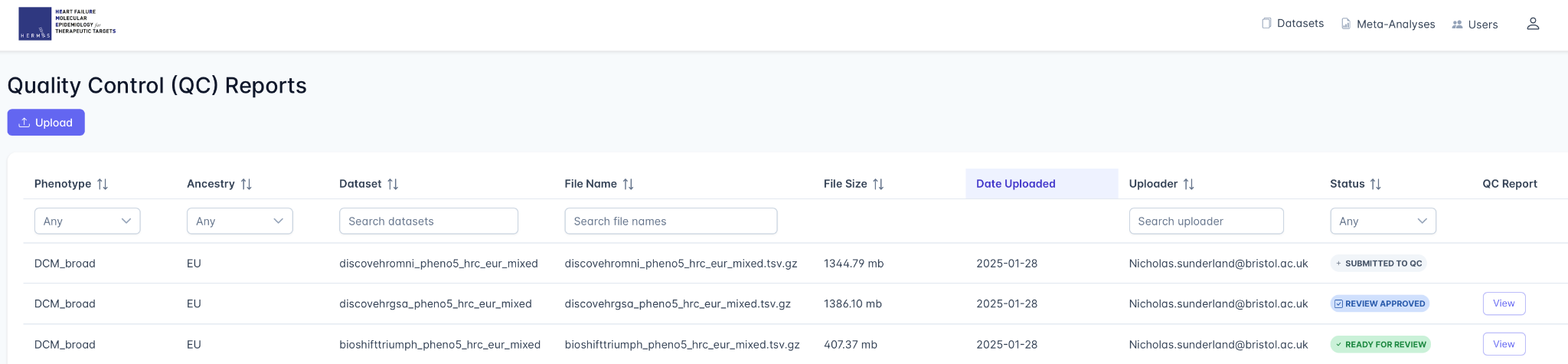

**Figure 2B - GWAS quality control study report**

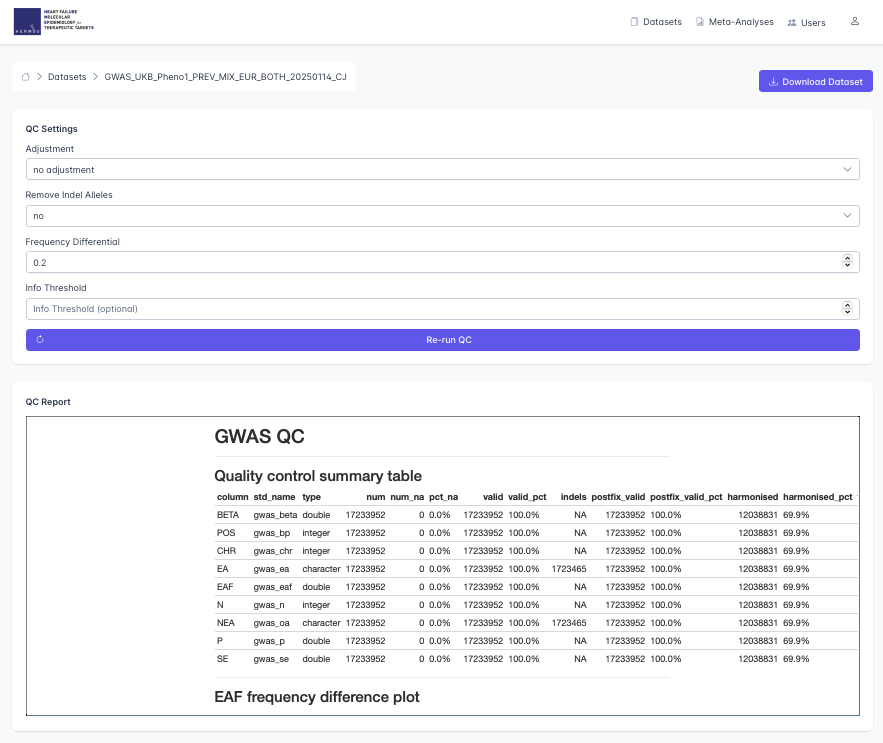

**Figure 2C - GWAS quality control plots**

| *A - allele frequency comparison plot*  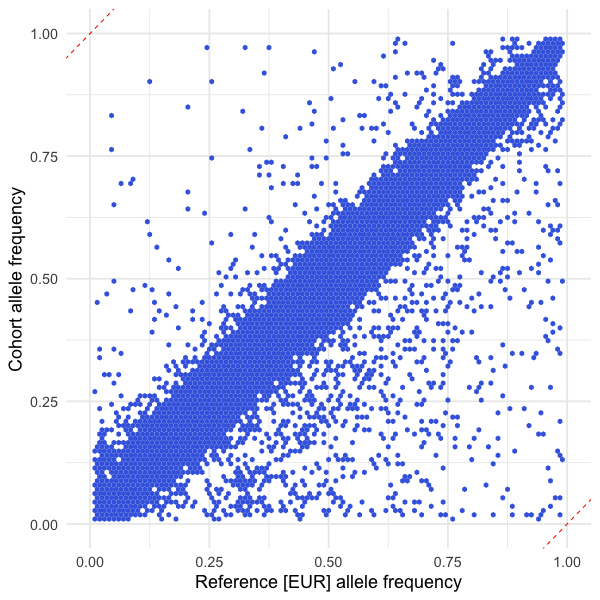 | B - QQ plot  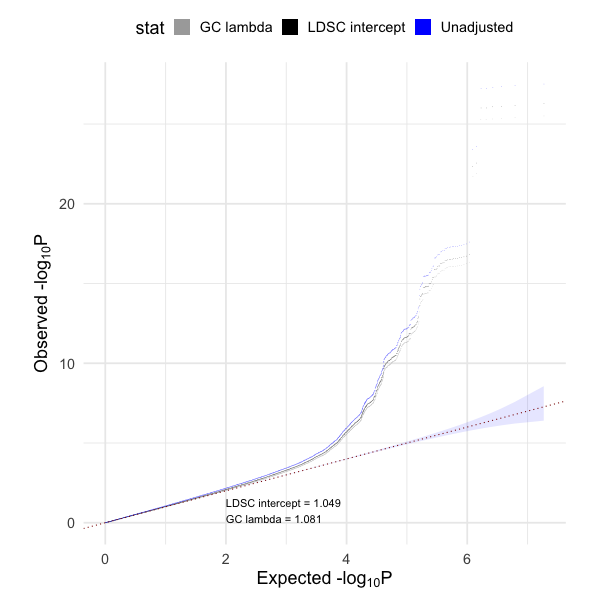 | C - PZ plot  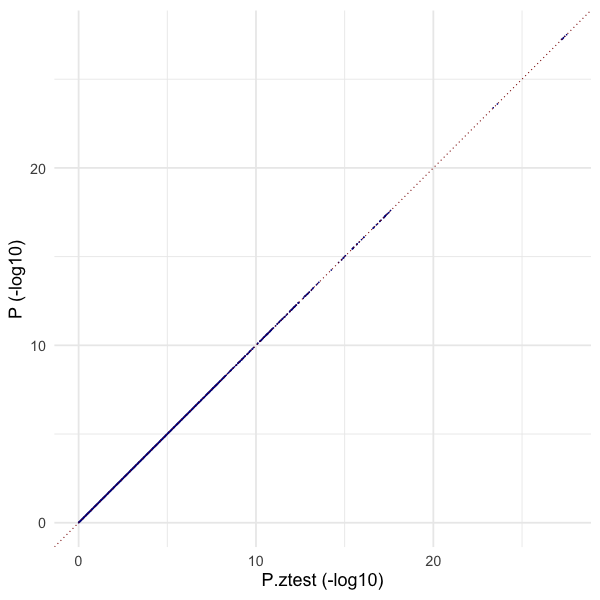 |
| --- | --- | --- |

**Figure 3A - Meta-analysis dashboard**

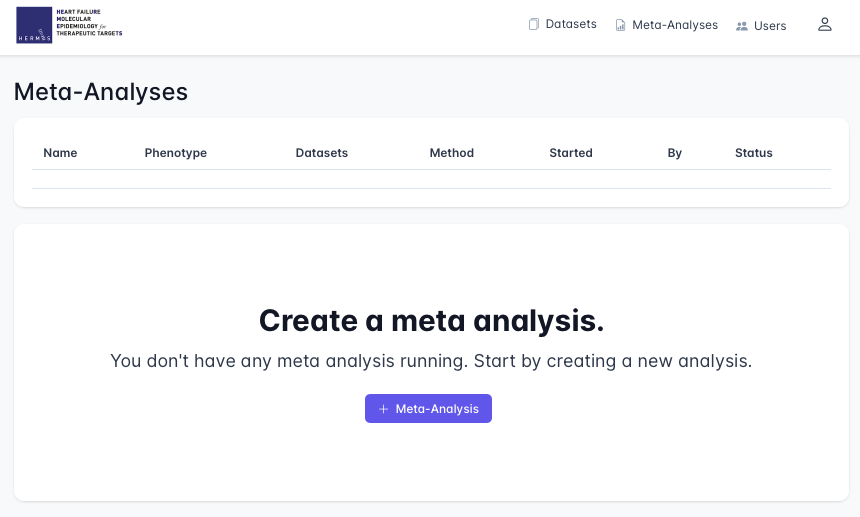

**Figure 3B - Meta-analysis report**

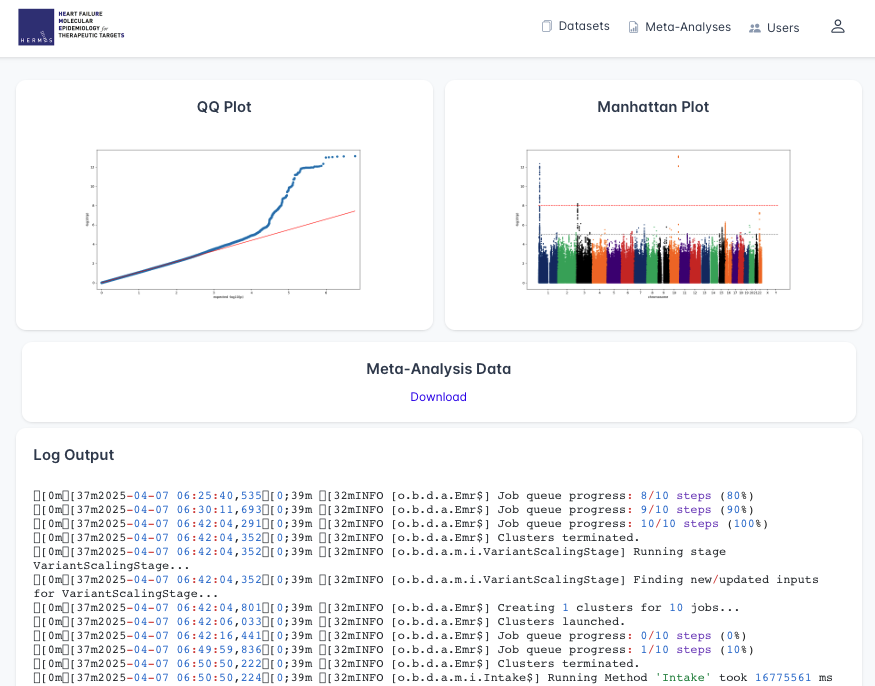

**Figure 4 - Dilated Cardiomyopathy GWAS Replication**

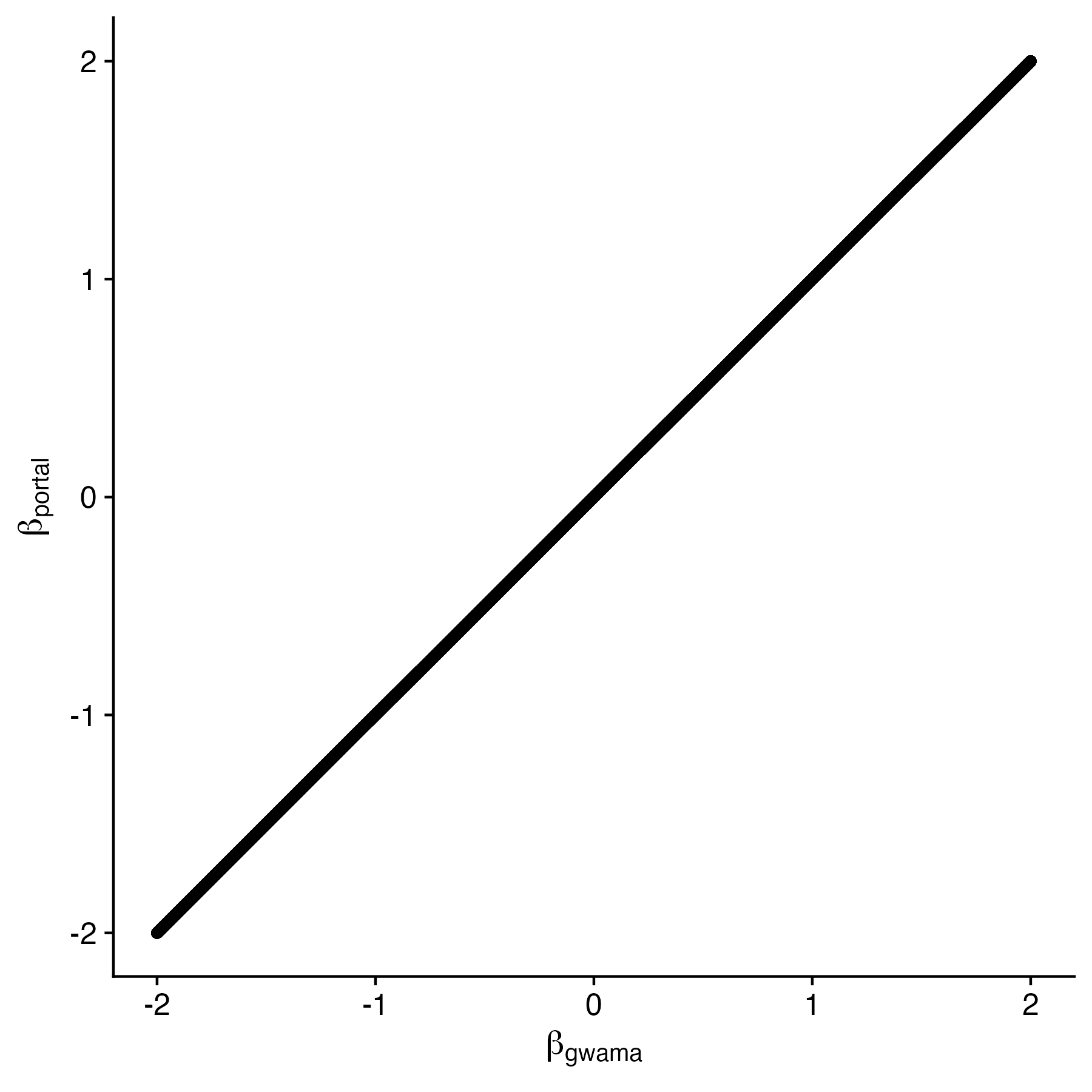
